# Evaluation of the safety and tolerability of three single ascending doses of Diamine oxidase (DAO) in healthy volunteers

**DOI:** 10.1101/2024.12.05.24318468

**Authors:** P. Molina Perelló, M. Puntes Rodríguez, J. Coimbra Hurtado, M. Garrido Sánchez, M. Castillo Ocaña, D. Martínez Bonifacio, L. Carrera Marcolin, J. Cuñé Castellana, R. Antonijoan Arbós

**Affiliations:** Centre d’Investigació de Medicaments (CIM-IR Sant Pau) Institut de Recerca Sant Pau, Barcelona, Spain; ABbiotek Health, Barcelona, Spain; Clinical Pharmacology departament, Hospital de la Santa Creu i Sant Pau, Barcelona, Spain

## Abstract

Diamine oxidase (DAO) is a key enzyme for metabolizing dietary histamine in the gastrointestinal tract. DAO deficiency can lead to histamine intolerance (HIT), manifesting as migraines, gastrointestinal disturbances, and allergic reactions. DAO supplementation has been shown to enhance histamine breakdown, alleviating these symptoms. This randomized, double-blind, single ascending dose (SAD) Phase I clinical trial aimed to evaluate the safety and tolerability of escalating doses of DAO supplementation in healthy volunteers. Thirty participants were enrolled and randomly assigned to receive DAO or placebo. Single doses of 42 mg, 84 mg, or 210 mg of DAO extract (adiDAO® Veg) were administered under fasting conditions. Vital signs, laboratory parameters, and adverse events (AEs) were monitored, and a follow-up visit assessed post-administration safety. All participants completed the study without discontinuations. No serious adverse events or clinically significant changes in vital signs, ECGs, or laboratory parameters were observed. The study demonstrated that even doses significantly exceeding typical recommendations were well-tolerated, with no safety concerns identified. This trial confirms the safety of high-dose DAO supplementation, supporting its potential use in managing DAO deficiency and HIT. Future studies are recommended to explore the effects of chronic high-dose administration and alternative dosage forms to improve convenience.

## Introduction

The diamino oxidase (DAO) enzyme plays a key role in breaking down histamine from many common foods in the gastrointestinal tract. Reduced DAO levels or impaired activity (DAO deficiency) can result in excessive unmetabolized histamine, which has been linked to various symptoms of histamine intolerance (HIT), such as migraines, gastrointestinal issues, and allergic processes [1]. DAO supplementation helps boost the enzyme’s activity in the small intestine, promoting local histamine breakdown and alleviating symptoms caused by food-related histamine intolerance. Histamine from the diet is mainly metabolized by DAO in the digestive system, and studies have shown that DAO supplementation can effectively treat histamine-induced intestinal issues [2,3].

Current recommended dose for DAO supplementation is 4.2mg of DAO extract, three times a day. This dose has shown clinical efficacy in reducing symptoms of histamine intolerance [3]. Although this is the recommended dose for DAO, in some cases, physiological and pathological processes may require higher doses of the extract.

A standard meal in the Mediterranean diet may contain up to 50 mg of histamine which does not cause any harm to nonsensitive patients. However, it is recognized by EFSA that, in a subgroup of the population, higher histamine content in the diet may cause undesirable reactions which may be due to lower activity of DAO, and which may be resolved with DAO supplementation [4].

Considering that endogenous DAO levels in some physiological situations, such as in pregnancy [5, 6, 7], can be greater than the recommended DAO oral dose, it is important to establish the safety and tolerability of the administered product indifferent ascending oral doses in healthy volunteers, as a first step to guarantee safety and tolerability of higher doses of DAO enzyme than the currently established.

### Objectives

The main aim of this study is to evaluate the safety and tolerability of the administered DAO product, of 3 different ascending doses. As the primary endpoint, safety and tolerability of the treatments will be evaluated by assessing vital signs, laboratory analyses, electrocardiogram (ECG) and incidence of adverse events (AE).

## Methods

### Ethics Approval

The study was approved by the local ethics committee and was performed in accordance with the Declaration of Helsinki. Written informed consent was obtained from all study participants. The study was performed at the Phase I Clinical Research Center, CIM-IR Sant Pau, Barcelona (Spain). Each subject was fully informed and provided consent to participate in the study, signing an informed consent form prior to performing any evaluation. A complete medical check-up was conducted in order to verify that the subject meets the inclusion criteria and none of the exclusion criteria.

### Study Design

This was a randomized, double blinded, single-center, single ascending dose (SAD) study to evaluate the safety and tolerability of 3 different doses of DAO. Three groups were included in this study, one for each dose.

### Treatment of study

Each mini-tablet contained 4.2 mg of adiDAO® Veg, pea (*Pisum sativum*) sprout dehydrated powder rich in DAO standardized at an enzymatic activity against histamine of >14.500 KHDU/g DAO (supplied by DRHealthcare España, S.L.). The following single-dose treatments were evaluated: 42 mg of DAO extract, once/day, 10 tablets with 240mL of water administration (Dose 1) or placebo; 84 mg of DAO extract, once/day, 20 tablets with 240mL of water administration (Dose 2) or placebo; 210 mg of DAO extract, once/day, 50 tablets with 240mL of water administration (Dose 3) or placebo. All administrations were performed under fasted conditions. The study product and placebo were randomly assigned using the program SPSS v. 26.0. At the beginning of study, subjects were allocated to a randomization number following a procedure of consecutive assignment.

All volunteers went to the centre the day before the product administration and were at site under surveillance, until +24h of the product administration.

### Participants

Included in the study were healthy male and female subjects aged 18-50 years, with negative drug screening tests and an agreement to refrain from any other drugs (including natural food supplements, vitamins and medicinal plants) within 14 days prior to taking the study supplement.

All healthy volunteers were determined to be healthy based on their medical history, physical examination results, virology testing (Hepatitis B, C and HIV) results, and routine laboratory haematology and biochemistry results before enrolment in the study.

### Safety Assessment

Vital signs, physical examination results, electrocardiograms (ECGs), safety laboratory variables, and adverse events (AEs) were monitored throughout the study. Laboratory measures included haematology, clinical chemistry, coagulation, and urinalysis were performed at screening visit and at End of Study (+24h).

Vital signs and ECG (systolic, diastolic blood pressure and heart rate) were performed at: baseline [pre-dose], [+1h], [+2h], [+4h], [+8h] and [+24h] post-administration. A follow up visit at 6-8 days after product intake was performed to ensure the safety of the volunteers.

### Statistics

The statistical analysis and data management was carried out at the CIM-IR Sant Pau. The analysis was performed using the program IBM-SPSS (v26.0). Statistical summaries were descriptive in nature. In general, data obtained in the trial are summarised descriptively and interpreted in an exploratory way. Descriptive statistics were provided by treatment group and scheduled visit. The default summary statistics for quantitative (continuous) variables are the number of observations (N), mean, standard deviation (SD), median, minimum (min) and maximum (max), for those subjects with data. For ordinal variable (t_max_), the number of observations (N), median and interquartile range, expressed as percentile 25 (P25) and percentile 75 (P75).

10 healthy volunteers (HVs) were enrolled in each dose level and were randomly assigned to DAO or placebo in an 8:2 ratio ratio (8 received active treatment, and 2 received the placebo). 30 volunteers were enrolled in total.

## Results

### Participant characteristics

All subjects randomised in the study met the inclusion criteria and none of the exclusion criteria at the beginning of the trial. The study was performed according to the procedures specified in the protocol. A total of 30 HVs (15 males and 15 females) were enrolled in the study. All 30 HVs received either a single dose of DAO or placebo and completed all the phases of the study. None of the HVs discontinued the study.

The characteristics of the observed HVs, including demographic data, are listed in Table 1.

**Table 1:** Baseline Demographics Abbreviations: BMI, body mass index; SD, Standard Deviation

|  | Mean | SD | Median | Min | Max | N |
| --- | --- | --- | --- | --- | --- | --- |
| Age (years) | 31.9 | 9.4 | 30.00 | 19.0 | 47.0 | 30 |
| Weight (kg) | 70.3 | 9.7 | 71.8 | 50.0 | 86.0 | 30 |
| Height (cm) | 168.9 | 9.6 | 170.5 | 151.0 | 190.0 | 30 |
| BMI (kg/m<sup>2</sup>) | 24.6 | 2.7 | 24.9 | 19.0 | 29.0 | 30 |

The randomized study population (N=30) was composed by Caucasian volunteers with a body weight within the normal range, without clinical evidence of laboratory, ECG or vital sign abnormalities. 26 of the randomised subjects were non-smokers and the remaining 4 subjects stopped smoking at least 6 months prior to the study treatment phase.

Regarding alcohol consumption, 18 subjects did not drink alcoholic beverages at all, and the remaining 12 subjects consumed a maximum of 10 g of alcohol per day (mean 3.26 g/daily). Regarding the xanthine’s consumption, 8 subjects participating in the study did not consume xanthines at all, and the remaining 22 subjects consumed xanthines (tea, coffee, chocolate, etc.) daily (mean 1.42 units/day). No subjects discontinued the study.

The median (range) values for the demographic characteristics of HVs who were enrolled in the study were as follows: 30 (19-47) years old, 170 (151-190) cm tall, 71.80 (50-86) kg, and a body mass index (BMI) of 24.9 (19-29) kg/m^2^.

No serious adverse events (SAEs) were recorded during the study. All participants tolerated the medication well. Safety examinations based on clinical and laboratory parameters (haematology and biochemistry) did not show clinically relevant changes from baseline. The changes observed in the vital signs (systolic and diastolic blood pressure and heart rate) did not show any clinically relevant abnormalities and all the parameters were within the normal range. No clinically significant changes were observed in the electrocardiogram recordings carried out during the study. Neither did the physical examinations show any clinically important changes. There were no deaths or other significant/serious adverse events during the study.

## Discussion

DAO extract is a vegetal ingredient obtained from pea sprout dehydrated powder. The extract is obtained by dehydration of pea sprouts. It is important to consider that DAO (diamine oxidase) is an enzyme naturally produced endogenously by the human body, and there are no established limits for DAO supplementation.

Reported animal studies (acute oral study and a 90-day sub-chronic toxicity study) in rats corroborate the safety of orally administered diamine oxidase of pig kidney source [8]. The acute study aimed to investigate oral toxicity of diamine oxidase at concentrations of either 0.5 ml or 5.0 ml DAO/kg body weight (bw) corresponding to the enzymatic activity of 52,500 and 525,000 U/kg bw, respectively. No signs of clinical toxicity were noted in any of the animals [8].

The sub-chronic study was conducted to investigate the toxicity potential of diamine oxidase preparation in rats following repeated oral exposure for over 90 days. In this study, Wistar rats (8/sex/group) were administered diamine oxidase preparation suspended in saline at doses of 0 (control), 80 (low), 400 (mid), and 800 (high) µL/kg bw/day via gavage for 90 days. The enzymatic activity and protein concentration of the diamine oxidase preparation was reported as 560,000 HDU (histamine degrading unit)/ml and 15.7 mg/ml, respectively [8].

The doses used in the *in vivo* study represent 100-, 500-, and 1000-fold increases of the amount of diamine oxidase present in the currently commercialized DRHealthcare product (12.6 mg of DAO extract). Based on findings from this study, the no observed adverse effect level (NOAEL) is determined as at least 800 µl/kg bw/day equivalent to 448,000 U/kg bw/day. Intake of 12.6 mg, in terms of diamine oxidase activity intake (HDU/kg bw/day), is at least 620 times lower than the NOAEL for a 70 kg adult.

In the present clinical study, we administer concentrations of 42 mg, 84 mg, and 210 mg, which are extremely lower than the NOAEL. Apart from the animal toxicity data, some clinical studies have already used doses of 25.2 mg of oral DAO daily without any record of adverse events. A clinical study in patients with migraine and cefalea used daily doses of 25.2 mg of pig kidney protein extract standardized at the same concentration of the adiDAO® Veg supplement tested in our study for a three-month period. No side effects were identified [9]. It is also published that during pregnancy the placenta secretes up to 1000 times more DAO than in non-pregnant periods [5, 7]. It is assumed to be a protection to the foetus. Not only no adverse events have been linked to that physiological increase but on the contrary, lack of capacity to increase DAO levels during pregnancy is associated with some disorders (pre-eclampsia) and miscarriage [6]. AdiDAO® Veg has been on the market in Europe as a dietary supplement since 2022 with recommended daily doses of 12,6 mg of the pea sprout dehydrated extract without any side effects reported so far.

In the current clinical study, thirty subjects were included in the safety population, 24 received active DAO formulation and 6 of them received placebo formulation. DAO enzyme supplementation appears safe and well-tolerated in the doses tested.

Based on the results obtained in our study adiDAO® Veg is a safe, well tolerated and valid alternative to pig kidney protein extract rich in DAO.

The product’s safety was thoroughly evaluated, and no adverse events were observed, even after increasing the dose by 50-fold. These findings suggest that the product is highly safe for its potential application for managing DAO deficiency in healthy individuals and those with diagnosed histamine intolerance.

A practical limitation of the study is the administration of a high number of capsules to test the active dose. Alternative formats, such as sachets, have been developed to allow for higher doses of DAO in a single administration, which could improve ease of intake in future studies. Additionally, this study did not assess the effects of chronic high-dose DAO supplementation, but the findings demonstrate that acute high-dose intake is safe and does not pose any risk to individuals. For future research, a longer study duration could provide further insights on the safety of DAO consumption.

## Conclusion

From the safety point of view, no relevant changes in vital signs, haematology or biochemistry were observed. No serious adverse events were recorded, and no pattern of abnormality was observed.

In view of these clinical results, they can be considered as solid points to justify the safety of DAO supplementation in healthy volunteers given over such a short period of time. This study aims to modulate the product’s dosage to target populations with more specific and higher requirements for the enzyme.

## Data Availability

All data produced in the present study are available upon reasonable request to the authors

